# Are text-message based programs targeting adolescents and their parents an acceptable approach to preventing adolescent e-cigarette use?

**DOI:** 10.1101/2024.04.18.24305994

**Authors:** Courtney Barnes, Lisa Janssen, Stephanie Mantach, Sam McCrabb, Heidi Turon, Daniel Groombridge, Kate Bartlem, Caitlin Bialek, Lucy Couper, Luke Wolfenden

## Abstract

Adolescent e-cigarette use (also known as vaping), globally and within Australia, has steadily increased in recent years, with e-cigarettes now the most heavily used nicotine-containing products amongst adolescents. In response to the rise in prevalence, governments have introduced legislation to curb the supply of e-cigarettes to adolescents. To supplement these legislation measures, leading (inter)national public health agencies have called for the development of education and communication programs aimed at reducing adolescent e-cigarette uptake. Text-message programs for adolescents represent a potentially potent approach to achieve this. In this study, we assessed the acceptability of text-messages sent to adolescents and their parents to target factors associated with e-cigarette use. Text-messages were found to be acceptable to both adolescents and their parents however the effectiveness of the program on adolescent e-cigarette use still needs to be established prior to broader scale-up and investment.

---

Adolescent e-cigarette use (also known as vaping), globally and within Australia, has steadily increased in recent years, with e-cigarettes now the most heavily used nicotine-containing products amongst adolescents.^(1)^ A recent report by the Australian Institute of Health and Welfare indicated that in 2019, only 9.6% of adolescents aged 14-17 years had tried e-cigarettes, with this figure nearly tripling to 28% in 2022–2023.^(1)^ A systematic review of global evidence concluded that serious adverse effects posed by e-cigarettes include acute lung injury, poisoning, burns and immediate toxicity through inhalation, including seizures.^(2)^ The review also concluded that amongst non-smoking adolescents, e-cigarettes provide no health benefits and double the odds of future tobacco use.^(2)^ The rise in youth e-cigarette use is a considerable concern to parents and caregivers. For example, surveys of Australian parents have reported that whilst 70-80% of parents were concerned that their child may try e-cigarettes, more than half of parents (57%) had never discussed e-cigarettes with their children.^(3)^ Given these concerns, the dramatic increase in prevalence and the strong link between adolescent e-cigarette use and subsequent initiation of tobacco use,^(2)^ preventing adolescent e-cigarette use is a public health priority as well as a potentially potent strategy to reduce nicotine-related cancer burden.

In recent years, governments internationally and within Australia have taken legislative action to limit the supply of e-cigarettes to adolescents. In 2024, the Australian government introduced a ban on the importation of all disposable e-cigarettes. This is to work in conjunction with legislation that prohibited the supply of e-cigarettes (both nicotine and those marketed as nicotine-free) to individuals under the age of 18 years.^(4)^ The World Health Organization and other (inter)national public health agencies recommend such legislative action to be supplemented with education and communication public health programs to enhance efforts to curb the emerging public health issue.^(5)^ Text-message interventions have proven to be an effective public health approach to improving other adolescent health behaviours, including tobacco use,^(6)^ and have the ability to be scalable and rapidly deployed through existing infrastructure. For example, services like NSW Health’s “Get Healthy” online health coaching service and school-based communication platforms already provide text-message programs for other health behaviours (e.g. nutrition and obesity prevention), and could be easily expanded to e-cigarettes.^(7)^

Whilst previous research has proven text-message programs to be effective in improving other health behaviours, and formative research indicates text-messages may also show promise for e-cigarettes,^(8)^ little is known about the acceptability of employing such an approach to prevent adolescent e-cigarette use amongst end-users (i.e. adolescents and their parents). Such information is needed prior to broader investment in the approach to determine if text-messages align with the values, preferences and needs of end-users, thus making it more likely for them to engage with such programs and lead to better outcomes. However, formative evaluations of current e-cigarette text-message programs have predominately focused on cessation and have targeted older age groups (e.g. university students).^(8)^ Promisingly, one prevention study conducted in the United States found that delivering text-messages focusing on the health harms of e-cigarettes were feasible and acceptable to adolescents aged 14-18 years.^(9)^ However, this study did not target or assess acceptability of the messages amongst parents, who play an integral role in influencing adolescent e-cigarette behaviours.^(10)^

As such, we aimed to explore the acceptability of text-messages, distributed to adolescents and their parents to target factors (i.e. barriers and enablers) associated with adolescent e-cigarette use. These theoretically informed text-messages were developed through a comprehensive co-design process, with factors identified through an extensive scoping review.^(10)^ The co-design process included focus groups, surveys and iterative text-message writing activities conducted with parents, adolescents, parenting research experts, Aboriginal and health promotion program managers, e-cigarette experts, and behavioural scientists to develop and refine the messages. Adolescent (aged 12-15 years) and parent dyads received a series of text-messages, delivered weekly over 12 weeks. Each message addressed a different factor(s) associated with adolescent e-cigarette use, including knowledge of harmful health effects, peer influence and social norms, refusal skills and the availability of parent support. To foster positive conversation amongst families, both the adolescent and parent text-messages targeted similar factors each week. However, the specific content of each text-message was tailored to each target group (i.e. adolescent or parent). Six months after receiving the first text-message, the acceptability of the text-messages was assessed via an online survey using a 5-point Likert scale (strongly disagree to strongly agree). Findings indicate that the text-messages were highly acceptable to both adolescents and their parents (Table 1). For example, 77% of adolescents and 94% of parents found the text-messages to be an acceptable way of receiving information about e-cigarettes; 86% of parents found the text-messages improved their ability to discuss e-cigarettes with their child; and 73% of adolescents and 91% of parents would recommend the program to others.

**Table 1.** Acceptability of the text-messages overall and by e-cigarette ever-use status, socioeconomic status and rurality.

|  | TOTAL | Ever-use |  | Socioeconomic status<sup>a</sup> |  | Rurality<sup>a</sup> |  |  |  |
| --- | --- | --- | --- | --- | --- | --- | --- | --- | --- |
|  |  | Yes | No | Low | High | Major cities | Inner regional | Outer regional | Remote/very regional |
|  | N (%) | N (%) | N (%) | N (%) | N (%) | N (%) | N (%) | N (%) | N (%) |
| **Adolescents** | N=30 | 7 (23) | 23 (77) | 10 (33) | 20 (67) | 25 (83) | 3 (10) | 2 (7) | 0 (0) |
| Text-messages are an acceptable method for receiving information about e-cigarettes. | 23 (77) | 2 (29) | 21 (91) | 9 (90) | 14 (70) | 19 (76) | 2 (67) | 2 (100) | 0 (0) |
| The text-messages improved my knowledge of the potential harms of e-cigarettes. | 23 (77) | 4 (57) | 19 (86) | 7 (78) | 16 (80) | 18 (75) | 3 (100) | 2 (100) | 0 (0) |
| Receiving the text-messages improved my ability to refuse e-cigarettes. | 18 (60) | 2 (29) | 16 (73) | 6 (67) | 12 (60) | 14 (58) | 2 (67) | 2 (100) | 0 (0) |
| I would recommend the text-message program to other adolescents. | 22 (73) | 3 (43) | 19 (83) | 8 (89) | 14 (70) | 17 (71) | 3 (100) | 2 (100) | 0 (0) |
| **Parents** | N=35 | 6 (17) | 29 (83) | 13 (37) | 22 (63) | 29 (83) | 6 (17) | 0 (0) | 0 (0) |
| Receiving text-messages is an acceptable method for receiving information about e-cigarettes. | 33 (94) | 6 (100) | 26 (90) | 13 (100) | 20 (91) | 28 (97) | 5 (80) | 0 (0) | 0 (0) |
| The text-messages were useful for improving my knowledge around e-cigarettes. | 31 (89) | 6 (100) | 25 (86) | 12 (92) | 19 (86) | 25 (86) | 6 (100) | 0 (0) | 0 (0) |
| Receiving the text-messages improved my ability to discuss e-cigarettes with my child. | 30 (86) | 5 (83) | 25 (86) | 10 (77) | 20 (91) | 24 (83) | 6 (100) | 0 (0) | 0 (0) |
| I would recommend the text-message program to other parents or guardians of adolescents. | 32 (91) | 6 (100) | 26 (90) | 12 (92) | 20 (91) | 27 (93) | 5 (83) | 0 (0) | 0 (0) |

<sup>a</sup>Participant postcodes were used to classify participants as being located in the least disadvantaged (high socioeconomic status) or most disadvantaged (low socioeconomic status) areas as per the 2021 Socio-Economic Indexes For Australia. Postcodes ranked in the top 50% of NSW were classified as least disadvantaged and the bottom 50% of postcodes as the most disadvantaged. Postcodes were also used to classify participant geolocation as 'major cities', 'inner regional', 'outer regional' or 'remote/very regional' based on the Accessibility/Remoteness Index of Australia.

Findings of the evaluation indicate that a text-message program targeting adolescents and their parents is an acceptable and promising approach to prevent adolescent e-cigarette uptake. However, the effectiveness of the program on adolescent e-cigarette behaviours needs to be established prior to broader scale-up and investment. As such, a factorial randomised controlled trial testing the potential effect of the text-messages on adolescent susceptibility to, and use of, e-cigarettes and tobacco is underway. Findings from this body of research will contribute to a currently limited evidence-base, and provide policymakers, practitioners and funders with guidance on the types of health promotion interventions targeting adolescents and their parents that are potentially effective in addressing this public health priority.

## Data Availability

All data produced in the present study are available upon reasonable request to the authors.

## Ethics approval statement

Ethics approval to conduct the research was obtained from the University of Newcastle Human Research Ethics Committee (H-2022-0340).

## Trial registration

This research was prospective registered with Australia New Zealand Clinical Trials Registry (ACTRN12623000079640).

## Acknowledgements

The authors thank the adolescents and parents who participated in the research project.

